# Deletion detection in SARS-CoV-2 genomes using multiplex-PCR sequencing from COVID-19 patients: elimination of false positives

**DOI:** 10.1101/2025.04.15.25325794

**Authors:** Nan Jiang, Colin N. Dewey, John Yin

## Abstract

Deletions are prevalent in the genomes of SARS-CoV-2 isolates from COVID-19 patients, but their roles in the severity, transmission, and persistence of disease are poorly understood. Millions of COVID-19 swab samples from patients have been sequenced and made available online, offering an unprecedented opportunity to study such deletions. Multiplex PCR-based amplicon sequencing (amplicon-seq) has been the most widely used method for sequencing clinical COVID-19 samples. However, existing bioinformatics methods applied to negative control samples sequenced by multiplex-PCR sequencing often yield large numbers of false-positive deletions. We found that these false positives commonly occur in short alignments, at low frequency and depth, and near primer-binding sites used for whole-genome amplification. To address this issue, we developed a filtering strategy, validated with positive control samples containing a known deletion. Our strategy accurately detected the known deletion and removed more than 99% of false positives. This method, applied to public COVID-19 swab data, revealed that deletions occurring independently of transcription regulatory sequences were about 20-fold less common than previously reported; however, they remain more frequent in symptomatic patients. Our optimized approach should enhance the reliability of SARS-CoV-2 deletion characterization from surveillance studies. Finally, our approach may guide the development of more reliable bioinformatics pipelines for genome sequence analyses of other viruses.

## Introduction

COVID-19 is caused by severe acute respiratory syndrome coronavirus 2 (SARS-CoV-2), a single-stranded positive-sense RNA virus. Globally, as of January 2025, there have been 777 million confirmed cases of COVID-19, including 7 million deaths, reported to the WHO (1). While extensive public health efforts and scientific advancements have helped mitigate the impact of the virus, SARS-CoV-2 continues to evolve, requiring ongoing genomic surveillance. Advances in sequencing technologies have enabled the collection of nearly 17 million publicly available SARS-CoV-2 genome sequences on GISAID (2). This wealth of data presents an unprecedented opportunity to study viral genome variants, including deletions, which can impact pathogenesis, transmission, and viral evolution (3, 4).

Deletion variants of SARS-CoV-2 can be broadly classified as replication-competent or defective. Replication-competent variants tend to have shorter deletions (less than 50 nucleotides) that remain in-frame, so deletions of 3*n* nucleotides translate into deletions of *n* amino acids (3, 5). Such variants have been found to confer adaptive advantages or disadvantages to the virus (3, 5, 6). Defective deletion variants, commonly called defective viral genomes (DVGs), lack at least one essential gene for growth and cannot reproduce in the absence of replication-competent virus. In general, DVGs can interfere with the viral genome replication and packaging, trigger antiviral immunity, and facilitate persistent infection (4). DVGs of SARS-CoV-2 have been detected in COVID-19 patients and virus cultures (7–10). Understanding the position, frequency, and distribution of deletion variants, both competent and defective, in clinical samples will likely be important for deciphering their roles in viral evolution and disease severity.

Due to the vast availability of COVID-19 sequencing data, large-scale analysis of SARS-CoV-2 deletions has become feasible. The most widely used sequencing method for clinical SARS-CoV-2 samples is multiplex-PCR amplicon sequencing (amplicon-seq) due to its cost-effectiveness and high sensitivity. In amplicon-seq, after reverse transcription, cDNAs are amplified by PCR using a set of primers designed from the known genome sequences in two reaction pools to generate a set of amplicons that tile the whole genome except for the 3’ and 5’ ends (11, 12). The ARTIC and Midnight protocols are two commonly used amplicon-seq approaches, differing in amplicon length and primer design (13–15).

Several tools have been developed to identify deletions from sequencing data. ViReMa (16), STAR (10), DI-tector (17), DVGFinder (18), and DVG-profiler (19) are designed to look for reads containing deletion junctions from Illumina short-read RNA-seq data. ViReMa and STAR are the most popular tools for detecting genome deletions. They find reads with parts that align to different parts of the genome. However, their application to amplicon-seq data has not been systematically validated. Unlike RNA-seq, amplicon-seq relies on targeted PCR amplification, which can introduce artifacts (20–22).

To improve the accuracy of deletion detection in amplicon-seq data, we systematically evaluated existing bioinformatics pipelines using negative and positive control samples. Our analysis revealed that false-positive deletions are common in short alignments, low-frequency reads, and regions near primer-binding sites. Based on these findings, we developed a filtering strategy that reduces false positives while preserving true deletions. We validated this strategy using synthetic control samples containing a known deletion, demonstrating that our method detects true deletions with minimal false positives. Using our refined pipeline on publicly available COVID-19 swab sequencing data, we found that deletions occurring independently of transcription regulatory sequences (TRS) are much less common than previously reported, but we confirm their higher prevalence in symptomatic individuals.

This study provides an improved bioinformatics framework for reliable deletion detection in SARS-CoV-2 multiplex-PCR sequencing data, and the approach can be readily adapted for studying other RNA viruses.

## Results

### 1. Existing methods predict many false positive deletions in negative controls

Various methods have been used to identify deletions in viral genomes from RNA-Seq data (7, 10), but their effectiveness in multiplex-PCR sequencing (amplicon-seq) remains untested. To address this gap, we evaluated representative pipelines from Gribble et al. (7) and Wong et al. (10) using Illumina multiplex-PCR sequencing data from negative controls in Kubik et al. (23). The main difference between these pipelines lies in their deletion detection software: Gribble et al. utilized ViReMa, while Wong et al. used STAR---both widely used for RNA virus analysis (7, 9, 10, 24, 25). Both pipelines generated numerous false positives (FPs) in negative controls (Figure 1). In a sample containing only human RNA (hRNA), ViReMa detected 691 unique FPs, while STAR identified 1,039. In a sample containing a mix of human RNA and synthetic SARS-CoV-2 genomic RNA (vgRNA), ViReMa and STAR detected 379 and 551 unique FPs, respectively. To further evaluate the impact of sequencing depth and viral genome copy number, we analyzed additional multiplex PCR sequencing data from control samples reported by Kubik et al., using both methods. We found that FP counts were not correlated with viral genome copy number but appeared to be influenced by sequencing depth (Supplementary Table 1).

**Figure 1.**
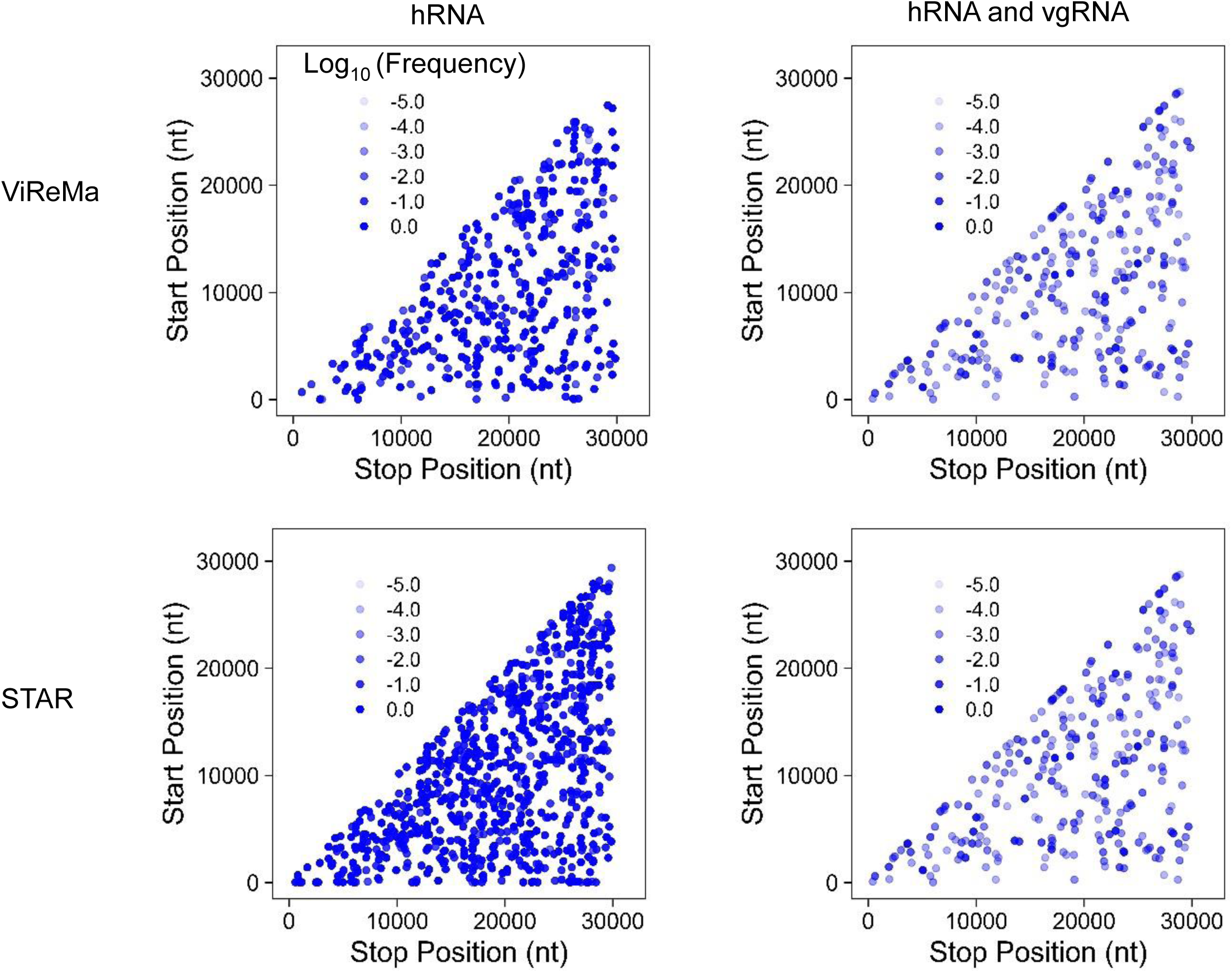
Prediction of false-positive (FP) deletion junctions by ViReMa (upper panels) and STAR (lower panels) from multiplex PCR Illumina sequencing. Deletion junctions in SARS-CoV-2 were identified from publicly available sequencing datasets derived from (i) human reference RNA (hRNA) only and (ii) a mixture of hRNA and synthetic SARS-CoV-2 viral genomic RNA (vgRNA). Deletion junctions are mapped according to their genomic position (5’ junction site, Start Position; 3’junction site, Stop Position). The transparency represents their log10(Frequency), so −2.0 has a frequency of 10^-2^ or 0.01.

To quantify the frequency of a specific deletion, we calculated the ratio of deletion depth (reads supporting the deletion) to the smaller of the depths at the deletion’s donor and acceptor sites (Figure 2). This method addresses uneven coverage in multiplex-PCR sequencing, where the smaller depth, representing the less efficient primer pair, influences deletion detection. In the hRNA-only sample, the low depth of the viral genome led to consistently high FP frequencies, whereas the mixture exhibited a broader distribution of FP frequencies (Figure 1).

**Figure 2.**
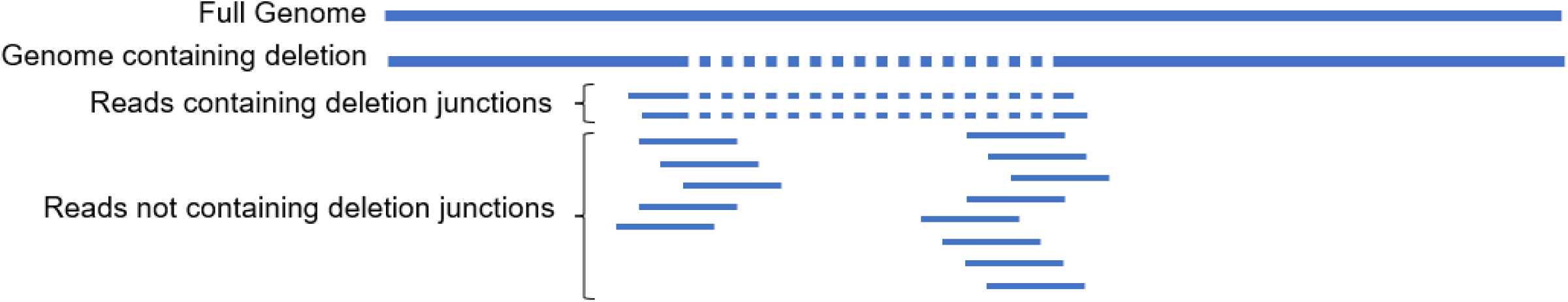
Calculation of deletion frequency. The specific deletion frequency is calculated using the number of reads containing a given deletion junction divided by the lower coverage depth between the start position and stop position of the deletion. Detection of a deletion junction requires amplification by primers flanking the start and stop sites; if either primer is missing, the deletion cannot be captured. The lower coverage depth reflects the less efficient primer set, which may limit detection sensitivity. In the example shown, the deletion frequency is 2/7 (approximately 0.29).

In addition to the publicly available sequencing data, we sequenced three further negative control samples using multiplex-PCR Illumina sequencing with the ARTIC v3 primer set: (1) vgRNA only, (2) hRNA only, and (3) a mixture of vgRNA and hRNA (vgRNA:hRNA = 1:300 by mass). The vgRNA and hRNA used were identical to those in the publicly available sequencing datasets (3). Consistent with the results from the public datasets, both pipelines predicted numerous false positives (FPs) in all three negative control samples (Table 1). In the absence of vgRNA, the low depth of the viral genome led to consistently high FP frequencies. Conversely, the presence of vgRNA in the mixture produced a broader range of FP frequencies.

**Table 1.**
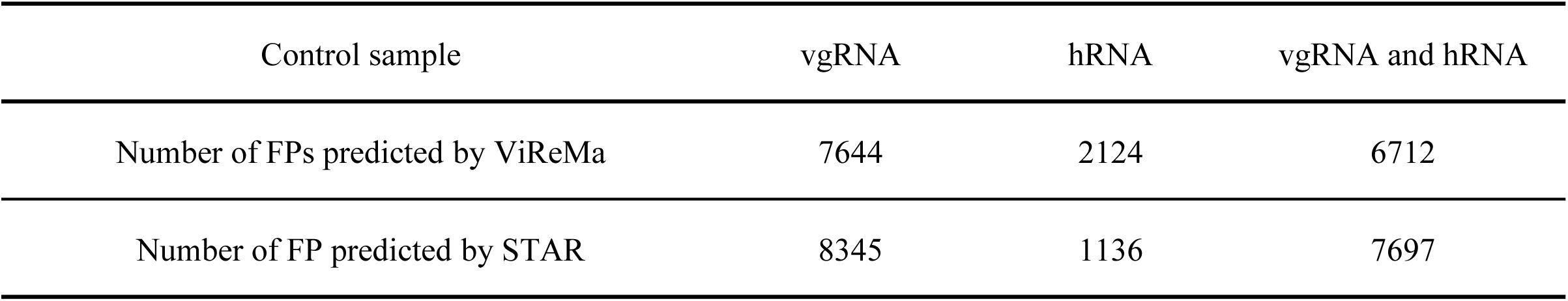
FPs in three negative control samples sequenced by multiplex-PCR Illumina sequencing using ARTIC v3 primer set before filtration.

### 2. False positive deletions often occur in short alignments and at low frequency

Analysis of the alignment files revealed that reads containing FPs could be stratified by the number of mapped nucleotides. In the hRNA sample, all FP reads had fewer than 75 mapped nucleotides, whereas the control sample containing both hRNA and vgRNA exhibited FP reads predominantly shorter than 75 nucleotides or longer than 100 nucleotides (Figure 3A). The hRNA sample lacked viral RNA, so SARS-CoV-2 sequences should be absent. Any alignments in the hRNA sample likely originated from primers used for cDNA amplification. Using our knowledge of the ARTIC v3 primer design, we examined reads containing FPs in the samples we sequenced. Analysis of the read alignments revealed that they clustered around ARTIC v3 primers. Plotting the distance between the start/stop positions of FP deletions and the nearest ARTIC v3 primer ends (5’ or 3’), showed that the average distance (3 nt) was significantly less than the average distance for random positions (41 nt) (Figure 3B). This finding suggests that most FPs were associated with ARTIC primers, potentially due to primer-dimers (26), especially in samples without vgRNA, in which most FPs came from reads with fewer than 75 mapped nucleotides (roughly twice the length of the ARTIC v3 primers).

**Figure 3.**
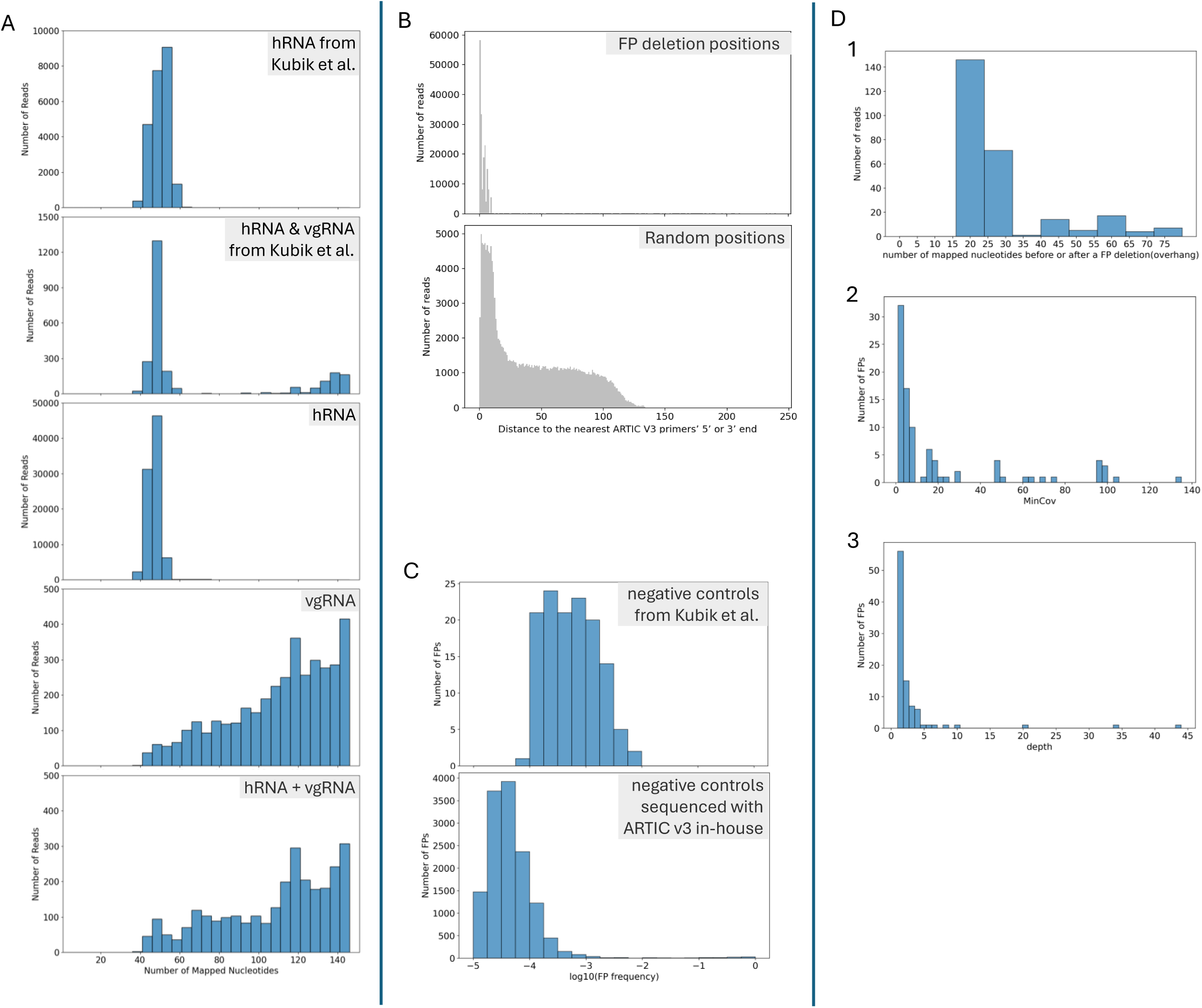
Characterization of FP deletions. (A) Distribution of the number of mapped nucleotides in FP-containing reads detected by ViReMa from samples of human reference RNA (hRNA), hRNA with synthetic viral genomic RNA (vgRNA) from Kubik et al., and hRNA, vgRNA, and hRNA + vgRNA samples sequenced using ARTIC v3 primers. (B) Distribution of distances of FPs’ start and end positions to the nearest ARTIC v3 primers’ 5’ or 3’end (upper panel), before filtration in hRNA sample sequenced by ARTIC v3 analyzed by ViReMa, For comparison, the lower panel shows the distribution of distances from randomly selected genomic positions (equal in number to the total FP start and stop positions) to the nearest primer ends. The average distances were 3 nt for FP deletion positions and 41 nt for random positions. (C) Distribution of FP frequency after removing reads with fewer than 75 nucleotides mapped to the reference genome, in the sequencing data of negative controls from Kubik et al. and all negative control samples sequenced with ARTIC v3 analyzed by ViReMa. (D) Analysis of FPs (mapped nucleotides ≥ 75 nts, frequency ≥ 0.01) detected by ViReMa in the three negative control samples (hRNA, vgRNA, and hRNA + vgRNA) sequenced with ARTIC v3. Shown are distributions of: (1) the smaller

Based on this characterization, we stratified the reads into two groups: (1) reads with fewer than 75 mapped nucleotides and (2) reads with at least 75 mapped nucleotides. In the data from Kubik et al., all FPs in the hRNA-only sample originated from read group 1. In contrast, for the vgRNA and hRNA mixture, 70% of the reads were classified as group 1, while 30% fell into group 2.

Most of the deletions in group 2 occurred at frequencies below 0.001, consistent with the typical error rate (∼0.001) observed in RNA virus amplicon sequencing (20). Similar patterns were observed in the negative control samples sequenced in our lab using the ARTIC v3 primer set. Most FPs in the hRNA-only sample originated from read group 1. In the vgRNA and hRNA mixture, deletions in read group 2 mostly had frequencies below 0.001 (Figure 3C).

Further characterization of high-frequency FPs in group 2 revealed additional features. Given that cDNA amplification occurs in two pools and library preparation involves random DNA fragmentation, true deletions should exhibit variability in overhang lengths (the number of mapped nucleotides before and after each deletion) across reads. However, most FPs lacked reads with both overhangs exceeding 30 nucleotides, suggesting that these short overhangs originated from primers (Figure 3D1). Moreover, due to the uneven sequencing depth across the genome, we observed that most FPs were predicted in regions with low coverage (≤20) (Figure 3D2) and were supported by a small number of reads (<5) (Figure 3D3).

### 3. Filtering eliminates most false positives while retaining known deletions

Based on the characteristics of FPs, we developed a series of filters to eliminate them. Reads with fewer than 75 mapped nucleotides were excluded, and only deletions with frequencies of at least 0.01 were retained. While FPs often had start or stop coordinates near primer sites, filtering out deletions with coordinates within 5 nt of primer sites would exclude 4,193 genomic positions (∼13% of the genome). Instead, we utilized deletion overhang lengths as a criterion. Given the maximum length of ARTIC v3 primers (30 nt), we applied a deletion-level filter requiring both the max_right_overhang and max_left_overhang to exceed 30 nt. Unlike read-level filters, this deletion-level approach ensures that a deletion interval is supported as long as some reads have longer overhangs, even if individual reads may have shorter overhangs.

In addition, we implemented filters based on sequencing depth and coverage. Deletions were considered only if they were supported by at least five reads (depth ≥ 5) and if both start and stop positions had coverage depths exceeding 20 (MinCov > 20). Furthermore, we required that deletions have both depth_positive and depth_negative greater than 2, as true RNA deletions should be detected in reads from both positive and negative strands after ARTIC-specific PCR amplification and library preparation.

In summary, our filters removed reads with fewer than 75 mapped nucleotides, and excluded deletions with fewer than 5 supporting reads, frequency below 0.01, ‘MinCov’ less than 21, ‘max_right_overhang’ or ‘max_left_overhang’ below 31, or either ‘depth_positive’ or ‘depth_negative’ less than 3 (Figure 4).

**Figure 4.**
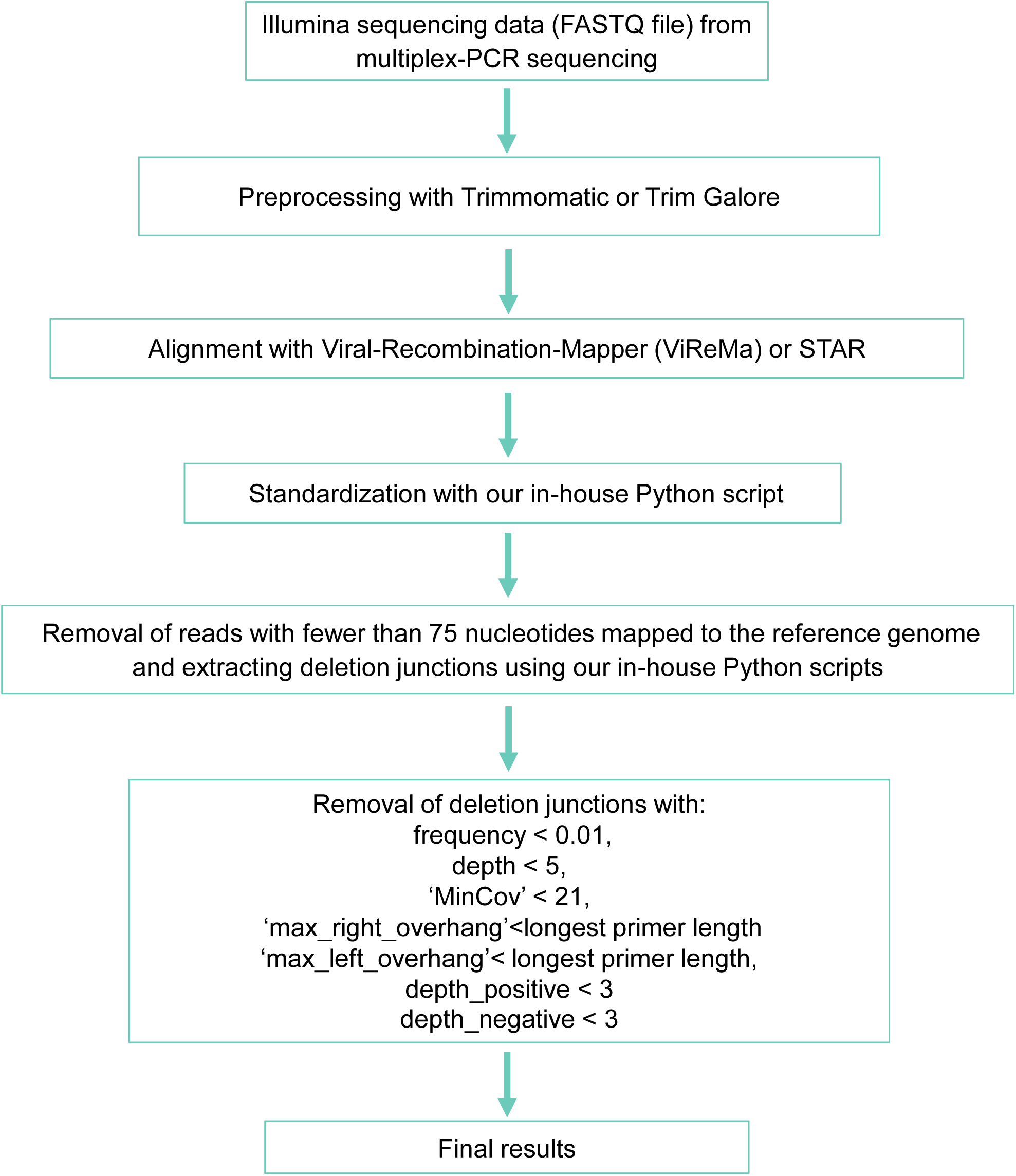
Optimized pipeline for identification of deletion junctions (>5 nt) in SARS-CoV-2 from multiplex-PCR Illumina sequencing.

To validate that the method could remove FPs while accurately detecting known deletions we used a synthetic deleted SARS-CoV-2 viral genome RNA (sDelVG) containing a deletion (10001–10689^26734–29903, inclusive coordinates), as shown in Figure 5 (9). Two types of positive control samples were prepared: sDelVG only, and a mixture of vgRNA, sDelVG, and hRNA (vgRNA:sDelVG:hRNA = 1:1:300 by mass). With the 1:1 ratio of vgRNA to sDelVG by mass, the expected frequency of the deletion was 0.91. The optimized filtering strategy eliminated most STAR FPs and faithfully detected the synthetic deletion in the positive control samples, while ViReMa did not predict any deletions after filtration (Table 2).

**Figure 5.**
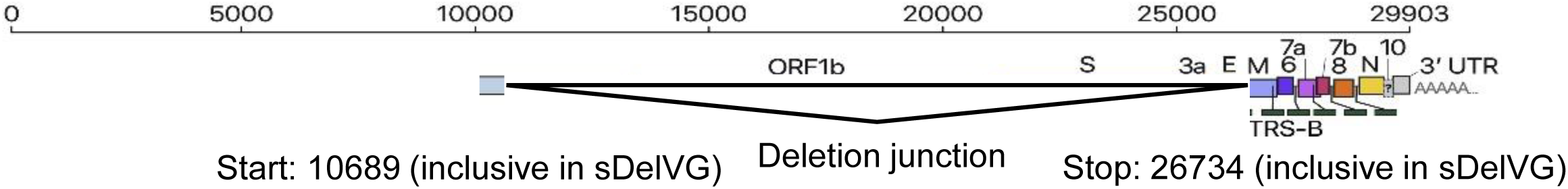
Synthetic deleted viral genome RNA (sDelVG) containing a deletion junction (10001-10689^26734-29903 (inclusive)) designed based on reference genome NC_045512.2. Due to the limitation of long RNA synthesis, we did not include the first 10000 nts of the SARS-CoV-2 genome in the sDelVG. The figure was adopted from Kim et al.(9).

**Table 2.** FPs and synthetic deletion (SD) in positive control samples sequenced by multiplex-PCR Illumina sequencing using ARTIC v3 primer set before and after the filters and standardization.

| Deletion detection tool | Positive control sample | sDeIVG | sDeIVG, vgRNA and hRNA |
| --- | --- | --- | --- |
| ViReMa | Number of FP before filtration | 2637 | 5147 |
|  | Number of FP after filtration | 0 | 0 |
|  | Number of FP after filtration and standardization | 6 | 0 |
|  | Frequency of the SD before filtration | 0.57 | 0.36 |
|  | Frequency of the SD after filtration | 0.00 | 0.00 |
|  | Frequency of the SD after filtration and standardization | 0.74 | 0.40 |
| STAR | Number of FP before filtration | 3940 | 5573 |
|  | Number of FP after filtration | 14 | 3 |
|  | Number of FP after filtration and standardization | 12 | 2 |
|  | Frequency of the SD before filtration | 0.93 | 0.55 |
|  | Frequency of the SD after filtration | 0.93 | 0.55 |
|  | Frequency of the SD after filtration and standardization | 0.91 | 0.55 |

Further examination of ViReMa’s deletion junctions before filtration revealed a deletion (10688^26733) near the synthetic deletion (10689^26734) in sDelVG. Notably, this deletion (10688^26733) appeared exclusively in negative-strand reads, while the deletion (10689^26734) was found only in positive-strand reads; these two predicted deletions likely originated from the synthetic deletion. However, both were removed by the filtering strategy because they were identified as two separate deletions that only existed on either positive or negative strand reads. Ideally, the sDelVG deletion should have been detected in both positive- and negative-strand reads after ARTIC cDNA amplification and library preparation. The coordinate discrepancy between strands for the same deletion came from the ambiguous alignment when there were nucleotide repeats on either side of the deletion junction. To address this issue, we developed a standardization process (standardize_alignments.py) for alignment files, ensuring each deletion was shifted as far downstream (3’ end) on the reference genome as possible, similar to the method that Martin et al. (24) used to fix a similar coordinate discrepancy problem. This standardization enabled ViReMa to detect the known deletion by consolidating deletions from different strand reads, which improved its estimates of deletion frequency but led to 6 FPs.

Currently, even with the latest alignment algorithms, it is challenging to predict deletion coordinates precisely when nucleotide repeats flank the junction. By combining filtration and standardization, we successfully detected the synthetic deletion in all the positive controls by both methods (Table 2) and eliminated all FPs in all the negative controls from both Kubik et al.’s and our sequencing data. Notably, standardization was unnecessary when using STAR, as it already handled the coordinate discrepancy between stands and produced higher and more accurate estimates of the synthetic deletion frequency than ViReMa.

To ensure the robustness of our filters with different primer sets, we re-sequenced the control samples using a different primer set, ARTIC v5. The filters effectively removed nearly all FPs in the control samples sequenced with ARTIC v5.3.2_400 (which includes primers up to 34 nt) and faithfully detected the synthetic deletion using both ViReMa and STAR (Table 3).

**Table 3.** FPs and SD in control samples sequenced by multiplex-PCR Illumina sequencing using ARTIC v5.3.2_400 primer set before and after the filters and standardization.

| Deletion<br>detection<br>tool | Control sample | vgRNA | hRNA | vgRNAand<br>hRNA | sDeIVG | sDeIVG,<br>vgRNA and<br>hRNA |
| --- | --- | --- | --- | --- | --- | --- |
| ViReMa | Number of FP before filtration<br>and standardization | 9297 | 3136 | 7804 | 3637 | 5486 |
|  | Number of FP after filtration<br>and standardization | 2 | 0 | 0 | 4 | 0 |
|  | Frequency of the SD before<br>filtration and standardization | 0.00 | 0.00 | 0.00 | 0.79 | 0.43 |
|  | Frequency of the SD after<br>filtration and standardization | 0.00 | 0.00 | 0.00 | 0.86 | 0.52 |
| STAR | Number of FP before filtration<br>and standardization | 9507 | 1112 | 8328 | 4233 | 6647 |
|  | Number of FP after filtration<br>and standardization | 3 | 0 | 0 | 8 | 0 |
|  | Frequency of the SD before<br>filtration and standardization | 0.00 | 0.00 | 0.00 | 0.94 | 0.59 |
|  | Frequency of the SD after<br>filtration and standardization | 0.00 | 0.00 | 0.00 | 0.94 | 0.59 |

We examined the estimated frequency of the synthetic deletion (10689^26734) in positive control samples (Tables 2 and 3). Both ARTIC v3 and ARTIC v5 allowed for detection of the deletion. But the amplification of vgRNA was more efficient than sDelVG with the multiplex primers designed to amplify the whole viral genome. Achieving a deletion frequency of one for sDelVG was challenging, as deletions located around the ends of reads were omitted; the deletion detection algorithm required a minimum number of aligned nucleotides both upstream and downstream of the deletion. Additionally, these ‘partially’ aligned reads were included in the coverage file, contributing to the lower observed deletion frequency. The differences in frequency using ARTIC v5 versus v3 primer sets are attributable to the different locations of the deletion on their respective amplicons. With ARTIC v3 primer sets, the deletion was primarily amplified by primers 36_LEFT (10666-10688) and 88_RIGHT (26890-26913), positioning the deletion at the 24th and 25th nucleotides from the 5’ end of the 204 bp amplicon. This resulted in many paired-end (2×150 bp) reads with the deletion near their ends, leading to false negatives. In contrast, with ARTIC v5 primer sets, the deletion was amplified by primers 35_LEFT (10527-10557) and 87_RIGHT (26989-27009), positioning it at the 163rd and 164th nucleotides of the 439 bp amplicon, allowing for better detection.

To further evaluate the ability of our filters to retain true deletions, we assessed whether they could detect canonical subgenomic mRNA (sgmRNA) deletions, which are commonly found in patient swab samples. We analyzed data from 81 patient swab samples sequenced with ARTIC v3 primers from Wong et al. (10) (Figure 6). Among the six sgmRNA deletions detectable in the majority of samples before filtering (S, E, M, 6, 7a, and N), four were reliably retained after applying our filters (S, M, 6, and N). To understand why some of the sgmRNA deletions were not detected before or after filtering, we calculated the minimum length of an amplicon needed to capture each deletion as well as the frequencies of those deletions that were detected. As the amplification efficiency of amplicons depends on the amplicon length, the detection of sgmRNA deletions related to not only the actual sgmRNA concentration but also the length of the amplicon containing the deletion. As expected, we found that the sgmRNA deletions corresponding to longer amplicons (3a, E, 7a, and 8) occurred at markedly lower frequencies and were thus often not detected before filtration or were filtered out. Further, although the amplicon for sgmRNA 7b is not long, it is known to occur at low abundance (9), and thus it was also generally filtered out.

**Figure 6.**
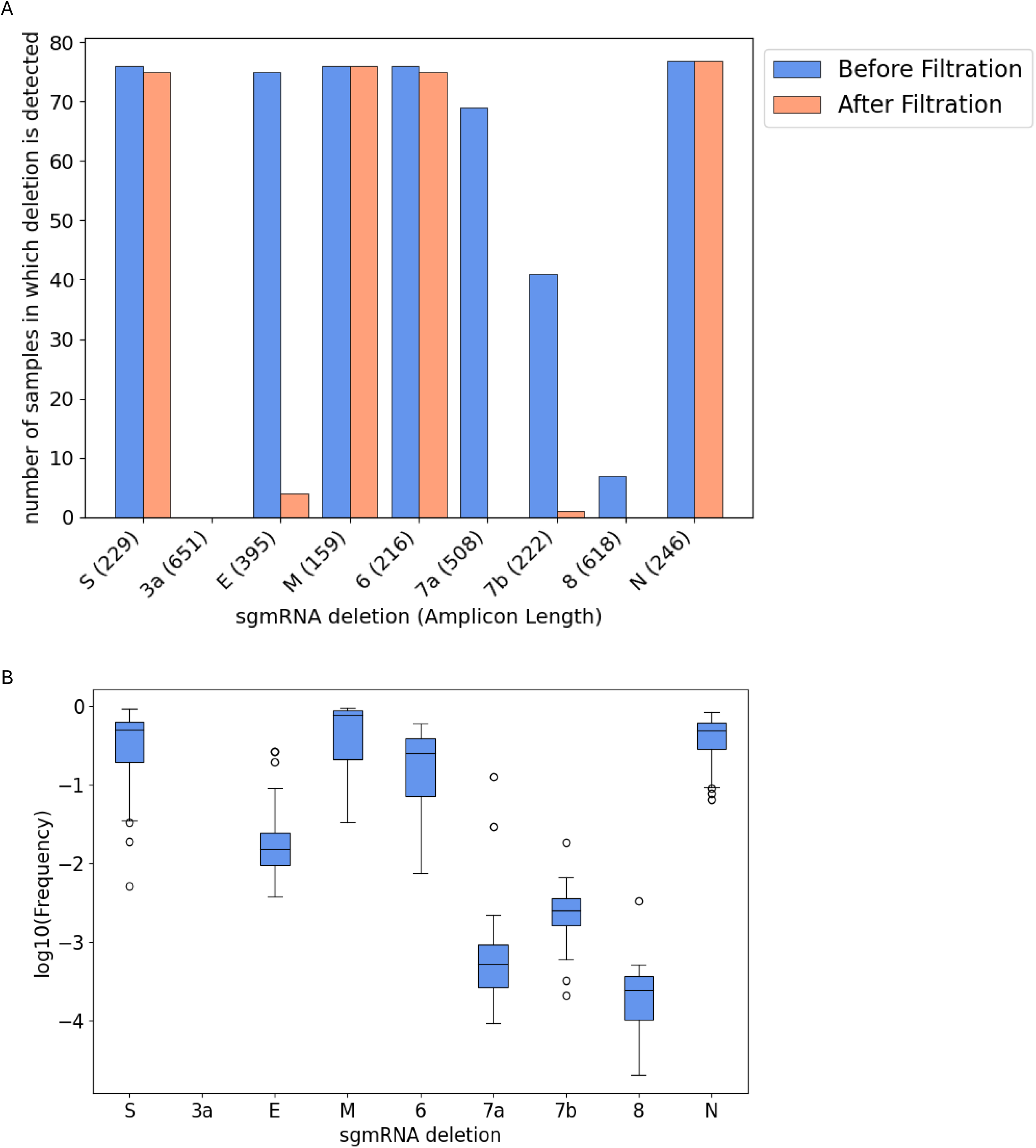
Detection of canonical SARS-CoV-2 subgenomic mRNA (sgmRNA) deletions in 81 COVID-19 swab samples. sgmRNA deletions were identified using STAR from multiplex PCR Illumina sequencing data of 81 swab

### 4. Application of the filtering strategy to COVID-19 swab data

Wong et al. sequenced 81 swab samples from patients (51 symptomatic, 30 asymptomatic) and analyzed them with STAR (10). They found that symptomatic hosts harbored more and longer TRS-independent deletions than asymptomatic hosts. To test the robustness of these results we initially repeated the published analysis and achieved similar results, to within 5 percent.

Specifically, Wong et al. found 501 total virus sequence deletions associated with patients who were symptomatic (463), asymptomatic (126), or both (88); using their approaches, we found 477 total deletions associated with patients who were symptomatic (441), asymptomatic (122), or both (86) (Figure 7A). However, when we analyzed the same data using our filtration and standardization approach we found more than 20-fold fewer deletions; specifically, we found only 22 total deletions associated with patients who were symptomatic (22), asymptomatic (1), or both (1) (Figure 7A).

**Figure 7.**
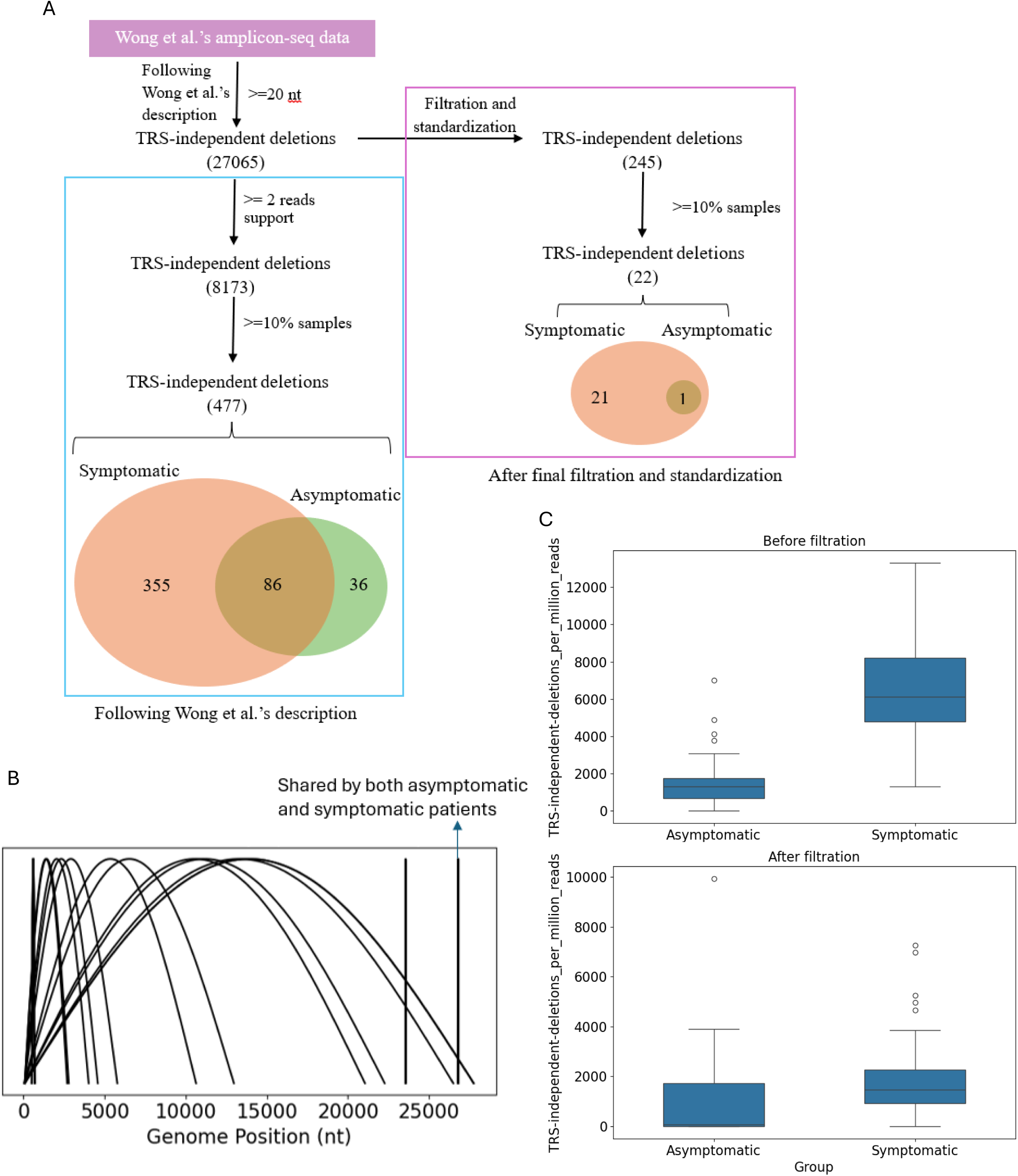
Filtration of COVID-19 swab sequencing data reduces the number of detected deletions but confirms their higher prevalence in symptomatic hosts. (A) TRS-independent deletions inferred by multiplex PCR Illumina sequencing data of specimens from asymptomatic and symptomatic patients. (B) Visualization of the TRS-independent deletions

Figure 7B highlights the genomic positions of the 22 deletions. Notably, all long deletions (>1000 nt) started within the first 100 nucleotides of the genome, suggesting that their formation is associated with subgenomic RNA generation. In contrast, only one short deletion (37 nt) was identified in ≥10% of asymptomatic patients.

We also analyzed the distributions of normalized TRS-independent split-aligned read counts in symptomatic and asymptomatic patients, before and after filtration and standardization. Although the number of TRS-independent split-aligned reads decreased after filtration, deletion events were more prevalent in viral samples from symptomatic hosts, consistent with Wong et al.’s conclusions (Figure 7C).

Finally, we evaluated the impact of various filters on the number of detected deletions (Table 4). The frequency filter (≥ 0.01) excluded the largest number of deletions, followed by the combined depth filter (depth ≥ 5 and depth_positive > 2 and depth_negative > 2).

**Table 4.** Effects of different filters on the number of TRS-independent deletions.

| Process (without secondary alignments) | Number of deletions |
| --- | --- |
| Without filtration and standardization | 27065 |
| Only with the number of aligned nucleotides on each read > 75 | 26936 |
| Only with standardization | 27071 |
| **Only with frequency >=0.01** | 418 |
| Only with depth >=5 and depth_positive > 2 and depth_negative > 2 | 3676 |
| Only with overhang length > 30 | 24795 |
| Only MinCov > 20 | 26941 |

## Discussion

The role of SARS-CoV-2 deletions in COVID-19 pathogenesis and viral evolution remains incompletely understood. The increasing availability of publicly shared sequencing data provides an unprecedented opportunity to investigate these deletions at a global scale. However, our study demonstrates that existing bioinformatics pipelines for deletion detection in multiplex-PCR sequencing data introduce a significant number of false positives (FPs).

By systematically evaluating negative control samples, we identified common characteristics of FP deletions, including their prevalence in short alignments, proximity to primer-binding sites, and low frequency. These insights allowed us to refine deletion detection strategies through a multi-step filtration process based on alignment length, deletion frequency, read depth, and overhang length.

Our optimized approach significantly improves the reliability of deletion detection by minimizing false positives while preserving true deletions. Through validation using synthetic deletion-positive controls, we confirmed that our method retains true positive deletions while eliminating erroneous calls.

Applying our refined filtration strategy to publicly available sequencing data revealed a substantially lower number of deletions than previously reported. For example, Wong et al. had originally identified thousands of TRS-independent deletions in patient-derived SARS-CoV-2 genomes. However, after filtering for false positives, we detected 20-fold fewer deletions, emphasizing the importance of stringent quality control measures in deletion detection. Despite the overall reduction in detected deletions, our results support the previous observation that symptomatic COVID-19 patients exhibit a higher prevalence of TRS-independent deletions compared to asymptomatic individuals.

While our optimized method effectively reduces false positives, several limitations remain in assessing the true positive rate. First, our analysis was constrained by the use of a single synthetic deleted viral genome (sDelVG) at only two concentrations. To ameliorate this issue, we also assessed the ability of the method to detect sgmRNAs, which may be considered known deletions in swab samples. The method reliably detects sgmRNA deletions with the exception of the sgmRNA deletions that required long amplicons and were found at low frequency.

Nevertheless, expanding validation experiments to include a broader range of deletion variants and concentrations would enhance the robustness of our approach, while also allowing for a more comprehensive exploration of the inherent tradeoff between false positive and true positive detections (precision versus recall) in methods like ours. Second, our frequency threshold of 0.01 prevents the detection of rare deletions, which may be relevant in studies focusing on low-frequency viral populations or emerging mutations. Third, while multiplex-PCR sequencing is the most widely used method for sequencing SARS-CoV-2 due to its high sensitivity and cost-effectiveness, it has intrinsic limitations in detecting deletions. Because this method relies on tiled amplicons for genome amplification in two reaction pools, deletions that begin before the 5’ end of the first primer in pool 2 or extend beyond the 3’ end of the last primer in pool 1 may not be detected. This limitation particularly affects the identification of sgmRNA deletions.

Additionally, multiplex-PCR sequencing cannot determine whether multiple distinct deletions originate from the same RNA molecule. Future work should explore complementary sequencing strategies, such as long-read sequencing or direct RNA sequencing, to overcome these limitations and the high input RNA requirement. Additionally, adapting our filtration approach to other RNA viruses could improve deletion detection across a broader range of viral genomic studies.

## Conclusion

Our study presents an optimized bioinformatics pipeline for reliable deletion detection in SARS-CoV-2 multiplex-PCR sequencing data. By systematically identifying and filtering out false positives, we provide a more accurate assessment of deletion prevalence in clinical samples. This refined approach enhances our ability to study defective viral genomes and their potential impact on viral evolution and disease progression. Despite the inherent limitations of multiplex-PCR sequencing, our method improves the reliability of deletion detection and can be readily adapted for other viral genomic studies. We hope that these findings will contribute to more accurate genomic surveillance efforts and inform future research on viral genome dynamics in SARS-CoV-2 and other RNA viruses.

## Materials and Methods

### 1. Multiplex-PCR Sequencing (Amplicon-Seq) of the Control Samples

Synthetic deleted SARS-CoV-2 viral RNA (sDVR) was ordered from Trilink. Synthetic SARS-CoV-2 genomic RNA (vgRNA) was purchased from Twist Bioscience (#102024). The human reference RNA was obtained from Invitrogen (QS0639). Control samples (Table 5) were reverse-transcribed using random primers to synthesize cDNA, which was amplified via two multiplex PCR reactions with ARTIC v3 or ARTIC v5.3.2_400 primers (IDT), following the NEBNext® ARTIC SARS-CoV-2 protocol <u>NEBNext ARTIC</u> <u>SARS-CoV-2 Library Prep Kit (Illumina) E7650 manual</u>. The primers were designed to tile amplicons across the viral genome. Amplified DNA was prepared using the QIAGEN FX DNA Library Preparation Kit (QIAGEN) with fragmentation, and sequenced with paired-end 150 bp reads on the Illumina NovaSeq X Plus platform.

**Table 5.** Control samples.

| Control Sample Components |
| --- |
| synthetic deleted SARS-CoV-2 viral RNA (sDVR) only |
| synthetic SARS-CoV-2 genome RNA (vgRNA) only |
| human reference RNA (hRNA) only |
| human reference RNA and synthetic SARS-CoV-2 genome RNA (gRNA: hRNA=1:300)* |
| human reference RNA, sDVR, and synthetic SARS-CoV-2 genome RNA (gRNA:sDVR:hRNA=1:1:300)* |

\*The ratios are calculated by mass.

### 2. Initial prediction of deletions with ViReMa

Initial prediction of deletions (>5 nt) with ViReMa followed the methodology of Gribble et al.(7) with modification. Detailed commands and parameters are provided in Supplementary methods. The workflow was as follows:

- Preprocessing: Raw reads were processed to remove Illumina TruSeq adapters using Trimmomatic (v0.39). Reads shorter than 75 bp were discarded, and low-quality bases (Q score < 30) were trimmed. Paired reads were renamed to append ‘_1’ and ‘_2’ for R1 and R2 reads, respectively, and concatenated into a single FASTQ file.
- Alignment: The concatenated FASTQ files were aligned to the SARS-CoV-2 reference genome (NCBI NC_045512.2) using ViReMa (Viral Recombination Mapper, v0.25).
- Depth and Frequency Calculation: Deletion coordinates were extracted from alignment file, including information on deletion position, deletion depth, overhang length, and aligned nucleotide count. The alignment file was processed using Samtools (v1.10) to calculate nucleotide depth (coverage) at each position. Deletion frequency was calculated as the depth of deletion divided by the smaller depth at the donor site and acceptor site.

### 3. Initial prediction of deletions with STAR

Initial prediction of deletions (>5 nt) with STAR followed Wong et al.(10) with modification. Detailed commands and parameters are provided in Supplementary methods. The method involved:

- Preprocessing: Raw paired-end reads were trimmed using Trim Galore (v0.4.3) via Cutadapt (v1.2.1). Reads shorter than 15 bp and low-quality bases (Q score < 30) were removed.
- Alignment: Trimmed reads were aligned to the SARS-CoV-2 reference genome (NCBI NC_045512.2) using STAR (v2.7.3a) with Wong et al.’s command set.
- Depth and Frequency Calculation: The deletion depth and frequency calculation methods were identical to those described in the ViReMa section.

### 4. Deletion summarization

Our final output file of deletion junctions contains the following information:

- chrom: reference name taken from FASTA file of mapped genome
- start: coordinate of the nucleotide right before the deletion (donor site)
- end: coordinate of the last deleted nucleotide (nucleotide right before acceptor site)
- depth: the number of reads containing the deletion
- depth_positive: the number of positive-strand reads containing the deletion
- depth_negative: the number of negative-strand reads containing the deletion
- n_fragments: the number of sequenced fragments (read pairs) containing the deletion
- max_left_overhang: the maximum number of mapped nucleotides before the deletion among reads containing the same deletion.
- max_right_overhang: the maximum number of mapped nucleotides after the deletion among reads containing the same deletion.
- max_aligned_length: the maximum number of mapped nucleotides among reads containing the same deletion
- start_cov: the coverage of the coordinate right before the deletion (donor site)
- end_cov: the coverage of the coordinate right after the deletion (acceptor site)
- MinCov: the lower coverage between the deletion’s start_cov versus end_cov
- Frequency: ‘depth’/ ‘MinCov’
- logFreq: log(‘Frequency’)
- deletion_length: length of the deletion (end-start)

### 5. Filtering strategy

The final optimized deletion identification pipeline is illustrated in Figure 4. Following the alignment of sequencing reads with ViReMa or STAR, the alignments were standardized using a Python script to shift deletions as far downstream (3’ end) as possible. Alignments with fewer than 75 mapped nucleotides were removed. Deletion coordinates were then extracted from the alignment file, and depth and frequency calculations were performed as described in the previous section. The final deletion summarization is provided above.

To remove likely false positives, deletion junctions that met any of the following conditions were excluded:

- Frequency < 0.01
- Depth < 5
- MinCov (minimum coverage at the deletion start or stop positions) < 21
- Maximum overhang on either end (max_right_overhang or max_left_overhang) < 31
- Depth of positive or negative strands < 3

### 6. Identification of deletions associated with transcription regulatory sequences (TRS-associated) and DVGs (TRS-independent)

We adopted the method from Wong et al. to distinguish between TRS-associated and TRS-independent deletion. We used the <u>translate.pl</u> script from Wong et al to translate the viral RNA genome containing a single deletion (assuming there is only one deletion per genome). If the translated peptide sequence matched any of the peptide sequences derived from subgenomic RNAs and the deletion’s start position was within the TRS-L (Transcription Regulatory Sequence-Leader) region, the deletion was classified as TRS-related. All other deletions were categorized as TRS-independent (DVG) deletions.

### 7. Canonical subgenomic RNA deletion identification

Since deletions were standardized by shifting them as far downstream (3’ end) as possible, the same approach was applied before identifying canonical subgenomic RNA (sgmRNA) deletions. To ensure consistency, we first standardized the coordinates of sgmRNA deletions on the SARS-CoV-2 reference genome (NCBI NC_045512.2) by shifting them to the 3’ end. The standardized coordinates of sgmRNA deletions are provided in Supplementary Table 2. A deletion was classified as an sgmRNA deletion only if its coordinates exactly matched those of the standardized canonical subgenomic RNA deletions.

## 8. Data and Code Availability

The multiplex-PCR sequencing data for negative control samples from Kubik et al. (23) were downloaded from NCBI (SRR13168458 and SRR13168423). Sequencing data for our control samples have been uploaded to NCBI (PRJNA1191883). COVID-19 swab sample sequencing data from Wong et al. were downloaded from NCBI (PRJNA690577). The code used for analysis is available on GitHub: https://github.com/NanJiang16/SARS-CoV-2_Deletion_detection_from_amplicon-seq The majority of analyses described here were run remotely with compute resources from the UW-Madison Center For High Throughput Computing (27).

## Data Availability

All data produced in the present work are contained in the manuscript. Specifically, the multiplex-PCR sequencing data for negative control samples from Kubik et al. were
downloaded from NCBI (SRR13168458 and SRR13168423). Sequencing data for our control
samples have been uploaded to NCBI (PRJNA1191883). COVID-19 swab sample sequencing
data from Wong et al. were downloaded from NCBI (SRA accession numbers in Supplementary
Table 1).

https://www.ncbi.nlm.nih.gov/bioproject/PRJNA681574/

https://www.ncbi.nlm.nih.gov/bioproject/?term=PRJNA690577

## Acknowledgments

We thank Dr. Peter Halfmann for his valuable support on this study. We are also grateful to the staff at the University of Wisconsin–Madison Biotechnology Center’s DNA Sequencing Facility (Research Resource Identifier – RRID:SCR_017759) for their technical assistance, and we acknowledge support from the 2022 Illumina Pilot Program (University of Wisconsin Biotechnology Center). We further acknowledge the computing resources and support provided by the UW–Madison Center for High Throughput Computing (CHTC), which were essential for the data analysis performed in this study.

This work was supported by the National Science Foundation (NSF) under grants DMS-2151959, MCB-2029281, and CBET-2030750; the National Institutes of Health (NIH) under awards OT2OD030524 and R01DK133605; and institutional support from the University of Wisconsin–Madison, including the Wisconsin Institute for Discovery, the Office of the Vice Chancellor for Research, and the Department of Chemical and Biological Engineering.

