## Supplemental Materials for "Deletion detection in SARS-CoV-2 genomes using multiplex-PCR sequencing from COVID-19 patients: elimination of false positives" (from another version of this preprint: version 1, 2025-04-16)

Supplementary Table 1. **FPs detected in control samples with different copy numbers of viral genomes before filtration with public available data from Kubik et al.** (SRR13168406, SRR13168373, SRR13168478, SRR13168434, SRR13168429)

| Copy number of viral genomes | 10 | 100 | 1000 | 10000 | 100000 |
|---|---|---|---|---|---|
| Number of FP predicted by ViReMa | 597 | 477 | 549 | 445 | 1355 |
| Number of FP predicted by STAR | 1270 | 1103 | 1196 | 829 | 1354 |
| Sequencing depth | 1.8M | 1.2M | 1.9M | 1.2M | 2.4M |

Supplementary Table 2. **Coordinates of standardized canonical subgenomic RNA (sgmRNA) deletions**

| sgmRNA | donor site | acceptor site |
|---|---|---|
| S | 75 | 21562 |
| 3a | 77 | 25393 |
| E | 77 | 26245 |
| M | 76 | 26480 |
| 6 | 75 | 27047 |
| 7a | 75 | 27394 |
| 7b | 79 | 27684 |
| 8 | 75 | 27894 |
| N | 75 | 28266 |

**Supplementary methods**

**ViReMa (v0.25)**

**With filtration and standardization:**

java -jar Trimmomatic-0.39/trimmomatic-0.39.jar PE ${name}_R1.fastq.gz ${name}_R2.fastq.gz output_${name}_paired_R1.fastq output_${name}_unpaired_R1.fastq output_${name}_paired_R2.fastq output_${name}_unpaired_R2.fastq ILLUMINACLIP:TruSeq3-PE.fa:2:30:10:2:True LEADING:3 TRAILING:3 SLIDINGWINDOW:4:15 MINLEN:75

python3 interleave_paired_end_fastq.py output_${name}_paired_R1.fastq output_${name}_paired_R2.fastq > interleaved_${name}.fastq

python3 ViReMa.py SARS-COV2_Reference_MN908947.3_padded.fasta interleaved_${name}.fastq ViReMa25_SARS2_${name}_recombinations.sam --Output_Dir ViReMa25_SARS2_${name} --Output_Tag ViReMa25_SARS2_${name} --Seed 20 -BED --MicroInDel_Length 5

python3 standardize_alignments.py SARS-COV2_Reference_MN908947.3_padded.fasta ./ViReMa25_SARS2_${name}/ViReMa25_SARS2_${name}_recombinations.sam > ./ViReMa25_SARS2_${name}/ViReMa25_SARS2_standardized_${name}_recombinations.bam

python3 filter_aligned_reads.py SARs-CoV-2_v5.3.2_400.primer.bed ./ViReMa25_SARS2_${name}/ViReMa25_SARS2_standardized_${name}_recombinations.bam --min-deletion-length 6 --max-overhang-primer-frac 1 --min-aligned-length 75 --virema > ./ViReMa25_SARS2_${name}/filtered_ViReMa25_SARS2_standardized_${name}_recombinations.bam

python3 extract_deletions.py ./ViReMa25_SARS2_${name}/filtered_ViReMa25_SARS2_standardized_${name}_recombinations.sorted.bam --primer-bed SARs-CoV-2_v5.3.2_400.primer.bed --min-deletion-length 6 --

virema > ./ViReMa25_SARS2_${name}/filtered_ViReMa25_SARS2_standardized_${name}_re
combinations.sorted.txt

samtools view -S -b ViReMa25_SARS2_${name}_recombinations.sam >
ViReMa25_SARS2_${name}_recombinations.bam

samtools sort ViReMa25_SARS2_${name}_recombinations.bam -o
ViReMa25_SARS2_${name}_recombinations.sorted.bam

samtools depth -a -m 0 ViReMa25_SARS2_${name}_recombinations.sorted.bam >
ViReMa25_SARS2_${name}_recombinations.coverage

**Without filtration and standardization:**

java -jar Trimmomatic-0.39/trimmomatic-0.39.jar PE ${name}_R1.fastq.gz
${name}_R2.fastq.gz output_${name}_paired_R1.fastq output_${name}_unpaired_R1.fastq
output_${name}_paired_R2.fastq output_${name}_unpaired_R2.fastq
ILLUMINACLIP:TruSeq3-PE.fa:2:30:10:2:True LEADING:3 TRAILING:3
SLIDINGWINDOW:4:15 MINLEN:65

python3 interleave_paired_end_fastq.py output_${name}_paired_R1.fastq
output_${name}_paired_R2.fastq > interleaved_${name}.fastq

python3 ViReMa.py SARS-COV2_Reference_MN908947.3_padded.fasta
interleaved_${name}.fastq ViReMa25_SARS2_${name}_recombinations.sam --Output_Dir
ViReMa25_SARS2_${name} --Output_Tag ViReMa25_SARS2_${name} --Seed 20 -BED --
MicroInDel_Length 5

samtools view -S -b ViReMa25_SARS2_${name}_recombinations.sam >
ViReMa25_SARS2_${name}_recombinations.bam

samtools sort ViReMa25_SARS2_${name}_recombinations.bam -o
ViReMa25_SARS2_${name}_recombinations.sorted.bam

samtools depth -a -m 0 ViReMa25_SARS2_${name}_recombinations.sorted.bam >
ViReMa25_SARS2_${name}_recombinations.coverage

python3
extract_deletions.py ./ViReMa25_SARS2_${name}/ViReMa25_SARS2_${name}_recombinatio
ns.sorted.bam --min-deletion-length 6 --

virema > ./ViReMa25_SARS2_${name}/annotated_ViReMa25_SARS2_${name}_recombinations.sorted.txt

**STAR (v2.7.3a)**

**With filtration and standardization:**

~/TrimGalore-0.4.3/trim_galore --stringency 3 -q 30 -e .10 --length 15 --paired ./${SRA}/${SRA}_1.fastq ./${SRA}/${SRA}_2.fastq

STAR --readFilesIn ./${SRA}_1_val_1.fq ./${SRA}_2_val_2.fq --outFileNamePrefix ${name} --genomeDir ./Genome_Dir --outFilterType BySJout --outFilterMultimapNmax 20 --alignSJoverhangMin 8 --alignSJDBoverhangMin 1 --outSJfilterOverhangMin 12 12 12 12 --outSJfilterCountUniqueMin 1 1 1 1 --outSJfilterCountTotalMin 1 1 1 1 --outSJfilterDistToOtherSJmin 0 0 0 0 --outFilterMismatchNmax 999 --outFilterMismatchNoverReadLmax 0.04 --scoreGapNoncan -4 --scoreGapATAC -4 --chimScoreJunctionNonGTAG 0 --chimOutType Junctions WithinBAM HardClip --alignSJstitchMismatchNmax -1 -1 -1 -1 --alignIntronMin 20 --alignIntronMax 1000000 --alignMatesGapMax 1000000

python3 standardize_alignments.py NC_045512.2.fasta ${name}Aligned.out.sam > ${name}_standardizationAligned.out.bam

python3 annotate_alignment_with_primers.py ARTIC_primers_v3.bed ${name}_standardizationAligned.out.bam > ${name}_annotated_standardization_Aligned.out.bam

python3 filter_aligned_reads.py ARTIC_primers_v3.bed ${name}_annotated_standardization_Aligned.out.bam --min-deletion-length 20 --max-overhang-primer-frac 1 --min-aligned-length 75 --primer-pool-matching --max-primer-dist 1 > filtered_${name}_annotated_standardizationAligned.out.bam

samtools sort filtered_${name}_annotated_standardizationAligned.out.bam -o filtered_${name}_annotated_standardizationAligned.out.sorted.bam

samtools depth -a -m 0 filtered_${name}_annotated_standardizationAligned.out.sorted.bam > filtered_${name}_annotated_standardizationAligned.out.sorted.coverage

python3 extract_deletions.py filtered_${name}_annotated_standardizationAligned.out.sorted.bam --primer-bed ARTIC_primers_v3.bed --min-deletion-length 20 --ignore-secondary > filtered_${name}_annotated_standardizationAligned.deletion.sorted.txt

**Without filtration and standardization:**

~/TrimGalore-0.4.3/trim_galore --stringency 3 -q 30 -e .10 --length 15 --
paired ./${SRA}/${SRA}_1.fastq ./${SRA}/${SRA}_2.fastq

STAR --readFilesIn ./${SRA}_1_val_1.fq ./${SRA}_2_val_2.fq --outFileNamePrefix ${name} -
-genomeDir ./Genome_Dir --outFilterType BySJout --outFilterMultimapNmax 20 --
alignSJoverhangMin 8 --alignSJDBoverhangMin 1 --outSJfilterOverhangMin 12 12 12 12 --
outSJfilterCountUniqueMin 1 1 1 1 --outSJfilterCountTotalMin 1 1 1 1 --
outSJfilterDistToOtherSJmin 0 0 0 0 --outFilterMismatchNmax 999 --
outFilterMismatchNoverReadLmax 0.04 --scoreGapNoncan -4 --scoreGapATAC -4 --
chimScoreJunctionNonGTAG 0 --chimOutType Junctions WithinBAM HardClip --
alignSJstitchMismatchNmax -1 -1 -1 -1 --alignIntronMin 20 --alignIntronMax 1000000 --
alignMatesGapMax 1000000

python3 annotate_alignment_with_primers.py ARTIC_primers_v3.bed
${name}Aligned.out.sam > ${name}_annotated_Aligned.out.bam

python3 filter_aligned_reads.py ARTIC_primers_v3.bed ${name}_annotated_Aligned.out.bam -
-min-deletion-length 20 --max-overhang-primer-frac 1 --primer-pool-matching --max-primer-dist
1 > ${name}_annotated_primer_Aligned.out.bam

samtools sort ${name}_annotated_primer_Aligned.out.bam -o
${name}_annotated_primer_Aligned.out.sorted.bam

samtools depth -a -m 0 ${name}_annotated_primer_Aligned.out.sorted.bam >
${name}_annotated_primer_Aligned.out.sorted.coverage

python3 extract_deletions.py ${name}_annotated_primer_Aligned.out.sorted.bam --primer-bed
ARTIC_primers_v3.bed --min-deletion-length 20 --ignore-secondary >
${name}_before_filteration_and_standardization_deletion.sorted.txt

**Identification of TRS-related and TRS-independent deletions**

tail -n +2 all_groups_all_deletions.txt | cut -f 1-2 | perl translate.pl 1>
all_groups_all_deletions_translated.xls 2> all_groups_all_deletions_translated.txt

(the deletion coordinates in the input of *translate.pl* are the first and last nucleotides deleted
(donor site+1, acceptor site -1))
